# Dengue in Cambodia 2002-2020: Cases, Characteristics and Capture by National Surveillance

**DOI:** 10.1101/2023.04.27.23289207

**Authors:** Christina Yek, Yimei Li, Andrea R. Pacheco, Chanthap Lon, Veasna Duong, Philippe Dussart, Sophana Chea, Sreyngim Lay, Somnang Man, Souv Kimsan, Chea Huch, Rithea Leang, Rekol Huy, Cara E. Brook, Jessica E. Manning

## Abstract

**Objective:** Data from 19 years of national dengue surveillance in Cambodia (2002-2020) were analyzed to describe trends in dengue case characteristics and incidence.

**Methods:** Generalized additive models were fitted to dengue case incidence and characteristics (mean age, case phenotype, fatality) over time. Dengue incidence in a pediatric cohort study (2018-2020) was compared to national data during the same period to evaluate disease under-estimation by national surveillance.

**Findings:** During 2002-2020, there were 353,270 cases of dengue (average age-adjusted incidence 1.75 cases/ 1,000 persons/ year) recorded in Cambodia, with an estimated 2.1-fold increase in case incidence between 2002 and 2020 (slope = 0.0058, SE = 0.0021, p = 0.006). Mean age of infected individuals increased from 5.8 years in 2002 to 9.1 years in 2020 (slope = 0.18, SE = 0.088, p <0.001); case fatality rates decreased from 1.77% in 2002 to 0.10% in 2020 (slope = - 0.16, SE = 0.0050, p <0.001). When compared to cohort data, national data under-estimated clinically apparent dengue case incidence by 5.0-fold (95% CI 0.2 – 26.5), and overall dengue case incidence (both apparent and inapparent cases) by 33.6-fold (range: 18.7-53.6).

**Conclusion:** Dengue incidence in Cambodia is increasing and disease is shifting to older pediatric populations. National surveillance continues to under-estimate case numbers. Future interventions should account for disease under-estimation and shifting demographics for scaling and to target appropriate age groups.

## Introduction

The global incidence of dengue is on the rise. Cases increased by 8-fold over the last two decades, with the greatest number of cases recorded in 2019.^1^ Experts attribute this trend to a variety of viral, vector, host, and ecologic factors including high rates of population growth in endemic areas, rapid unplanned urbanization without simultaneous expansion of vector control and water management systems, increasing global connectivity, and climate change expanding the geographic range of the *Aedes spp*. mosquito, among others.^2^

Accurate estimates of dengue burden are crucial to ensure appropriate allocation of resources to contain infection and manage clinical disease. The development of novel quadrivalent vaccines has added urgency to the task as public health entities plan vaccine rollouts in dengue-endemic areas.^3–5^ However, the most heavily-afflicted countries rely on resource-scarce national surveillance systems with predominantly clinicosyndromic case identification and passive reporting, leading to gross under-estimation of disease burden.^6^ One approach to this issue involves drawing data from active cohort surveillance and applying an expansion factor (i.e., the ratio of detected dengue cases to those in official reports) to estimate true incidence within the overall population.^6^ However, most active surveillance studies rely on the detection of symptomatic cases at select diagnostic centers, overlooking the majority of people that may have clinically inapparent infection or do not seek care for other reasons.^7^ Capturing this population of inapparent cases is vital to understand disease pathogenesis and host immunity, and importantly, for design and evaluation of containment strategies.^8^ Cross-sectional serosurveillance of dengue captures prior infection, both clinically apparent and inapparent. Where age-stratified data is available, this can be used to calculate transmission intensity (i.e., force of infection, defined as the rate at which susceptible individuals become infected) with practical applications including defining target populations for pre-vaccination serologic screening.^9^ Serosurveillance studies applying techniques such as plaque reduction neutralization assays (PRNT) can provide serotype-specific granularity and are less subject to cross-reactivity with other endemic flaviviruses than ELISA-based serologic assays.^10^ Finally, longitudinal serosurveillance provides the most accurate capture by removing assumptions about incidence rate constancy over time or age group. However, such methods are resource-intensive and impractical to guide public policy at a national level.^11^

The World Health Organization (WHO) region of Southeast Asia (SEA) includes ten dengue-endemic countries that comprise 70% of dengue cases worldwide.^1^ Among these is Cambodia, a country in the Indochinese peninsula with a population of 17 million. National dengue surveillance data for Cambodia was last published in 2010.^12^ This report covered a period of dynamic change from 1980 to 2007 during which post-civil war improvements in public health infrastructure led to improved surveillance efforts including several sentinel sites in the early 2000s. Since the publication of this report, Cambodia has continued to undergo marked changes in population demographics and urbanization. However, national dengue surveillance continues to have several limitations including reliance on self-referral, predominant clinicosyndromic identification, exclusion of private sector practices, and a largely passive reporting system dependent on variable participation of provincial facilities. The true incidence of disease is significantly under-estimated by national data: two studies conducted in central Cambodia between 2006 and 2008 extrapolated expansion factors from single-province active surveillance and described under-estimation of total dengue cases by national surveillance methods ranging from 3.9- to 29.0-fold.^13,14^ With novel interventions and vaccines on the horizon, it is paramount to improve estimates of dengue burden in countries such as Cambodia to better guide containment strategies and vaccine rollout campaigns.

In this report we analyze nearly two decades of Cambodian national dengue surveillance data from 2002 through 2020 and compare findings to those derived from a longitudinal pediatric cohort study to describe recent trends in dengue case characteristics and incidence.

## Methods

### National Surveillance for Dengue Cases

The Cambodia National Dengue Control Program collects monthly data on hospitalized dengue cases at public healthcare facilities. WHO clinical case definitions are applied to make a clinical diagnosis of dengue fever (DF), dengue hemorrhagic fever (DHF), and dengue shock syndrome (DSS). Serologic confirmatory testing is performed in a minority of cases using the SD BIOLINE Dengue Duo rapid test (Abbott®), depending on the patient ‘s ability to afford the test and the availability of test kits. In addition, the Virology Unit at Institut Pasteur du Cambodge confirms dengue serotype using a specific RT-PCR in a subset of patient samples submitted by sentinel sites.^15^ Sentinel surveillance was introduced in 5 provinces in 2001, with subsequent expansion to 10 provinces in 2020 and 15 provinces in 2021. Patients are treated according to the Cambodian National Guidelines for Clinical Management of Dengue 2018.^16^

### Census Data

Census data was obtained from the 1998, 2008, and 2019 national Cambodian population census. Population growth rate was computed using the exponential growth formula to extrapolate data for inter-census years.^17^

### Pediatric Cohort Study

A prospective longitudinal cohort study of 771 children aged two to nine years was conducted in the province of Kampong Speu, Cambodia to investigate the relationship between dengue infection and host immunological response to *Ae. aegypti* mosquito saliva (NCT03534245).^18^ Children were followed semi-annually from July 2018 through September 2021 with serial serologic testing using dengue IgG ELISA (PanBio Dengue Indirect IgG, Abbott Laboratories) and PRNT, as previously described.^18^ Children who developed symptoms consistent with acute dengue were evaluated as unscheduled sick visits, during which rapid dengue testing (SD Bioline DengueDuo NS1 Ag/immunoglobulin M [IgM]/IgG) was performed followed by confirmatory dengue PCR. Clinically inapparent cases of dengue were identified, defined as children who had negative dengue IgG ELISA at baseline and developed mono- or multitypic immunity (PRNT_50_ >1:40) during the study period. There was moderate loss to follow-up from mid-2020 onwards as a result of the global COVID-19 pandemic; due to paucity of data during this period, data was censored at the 4^th^ study visit in April 2020.

### Study Oversight and Ethics Statement

National dengue surveillance data used in this study were de-identified and are publicly available by request from the Cambodian Ministry of Health. Monthly dengue incidence is also accessible via WHO weekly reports.^19^ The U.S. National Institutes of Health and the National Ethics Committee on Human Research in Cambodia gave ethical approval for this work (pediatric protocol NCT03534245).

### Statistical Analysis

Crude dengue case data were collected from January 2002 to December 2020. Overall case and age-specific incidence rates were calculated using population data derived from 1998, 2008, and 2019 census (as above). Age-adjusted incidence was calculated by multiplying age-specific incidence and weighting of each age strata derived from 1998 census data.^12^ Longitudinal trends in dengue incidence, demographics, and case characteristics were explored using generalized additive models fitted to annual case data, with a monthly smoothing term (Supplemental Text S1). Two expansion factors were calculated by comparing dengue incidence in the pediatric cohort to that reported via national surveillance. Expansion factor 1 (*EF*_*1*_) represented under-detection of clinically apparent dengue and was calculated by determining the average incidence rate ratio of apparent dengue cases detected through active febrile surveillance in the pediatric cohort (*P*_*A*_) to that detected by national surveillance during July 2018-April 2020 for the same age group (children aged 2-9 years) living in Kampong Speu (*P*_*N*_). This was performed by fitting a Poisson generalized linear model via quasilikelihood to allow for variance inflation, adjusting for month as a categorical variable. Expansion factor 2 (*EF*_*2*_) represented under-detection of both clinically apparent and inapparent dengue and was calculated by dividing the cumulative incidence of apparent and inapparent dengue cases detected at semi-annual surveillance timepoints in pediatric cohort (*P*_*T*_) by semi-annual cumulative incidence of dengue cases in the corresponding age group in Kampong Speu detected by national surveillance (*P*_*N6*_). Statistical analyses were performed in R (version 4.1.0).

## Results

### Nationally Reported Dengue Incidence in Cambodia 2002-2020

Cases of dengue across 25 Cambodian provinces were reported on a weekly basis and collated in a central database. Over 19 years of national surveillance, a total of 353,270 cases were reported, corresponding to an average age-adjusted incidence of 1.75 cases/ 1,000 persons/year (Table 1). Major epidemics occurred in 2007, 2012, and 2019 (Figure 1A). The 2019 epidemic was the largest since national surveillance was implemented in 1980, with an annual total of 68,597 cases (age-adjusted incidence 6.27 cases/ 1,000 persons/year), a 3.91-fold increase from the average across prior years and a 1.64-fold increase from the average in other epidemic years (2007, 2012). Annual peaks in dengue cases occurred in June through August, coinciding with the country ‘s annual wet season and periods of heavy rainfall (Figure 1B).^20^ Siem Reap and Phnom Penh provinces reported the highest case incidence (average 3.00 and 2.03 cases/ 1,000 persons/year, respectively) (Supplementary Figure 1). Generalized additive models were fitted to dengue cases from 2002 to 2020 and demonstrated a significant increase in both crude case numbers (slope = 0.042, SE = 0.00031, p <0.001) and age-adjusted incidence (slope = 0.0058, SE = 0.0021, p = 0.006), representing a 2.1-fold increase in both metrics over the 19-year period (Figure 2 and Supplementary Table 1).

**Table 1:**
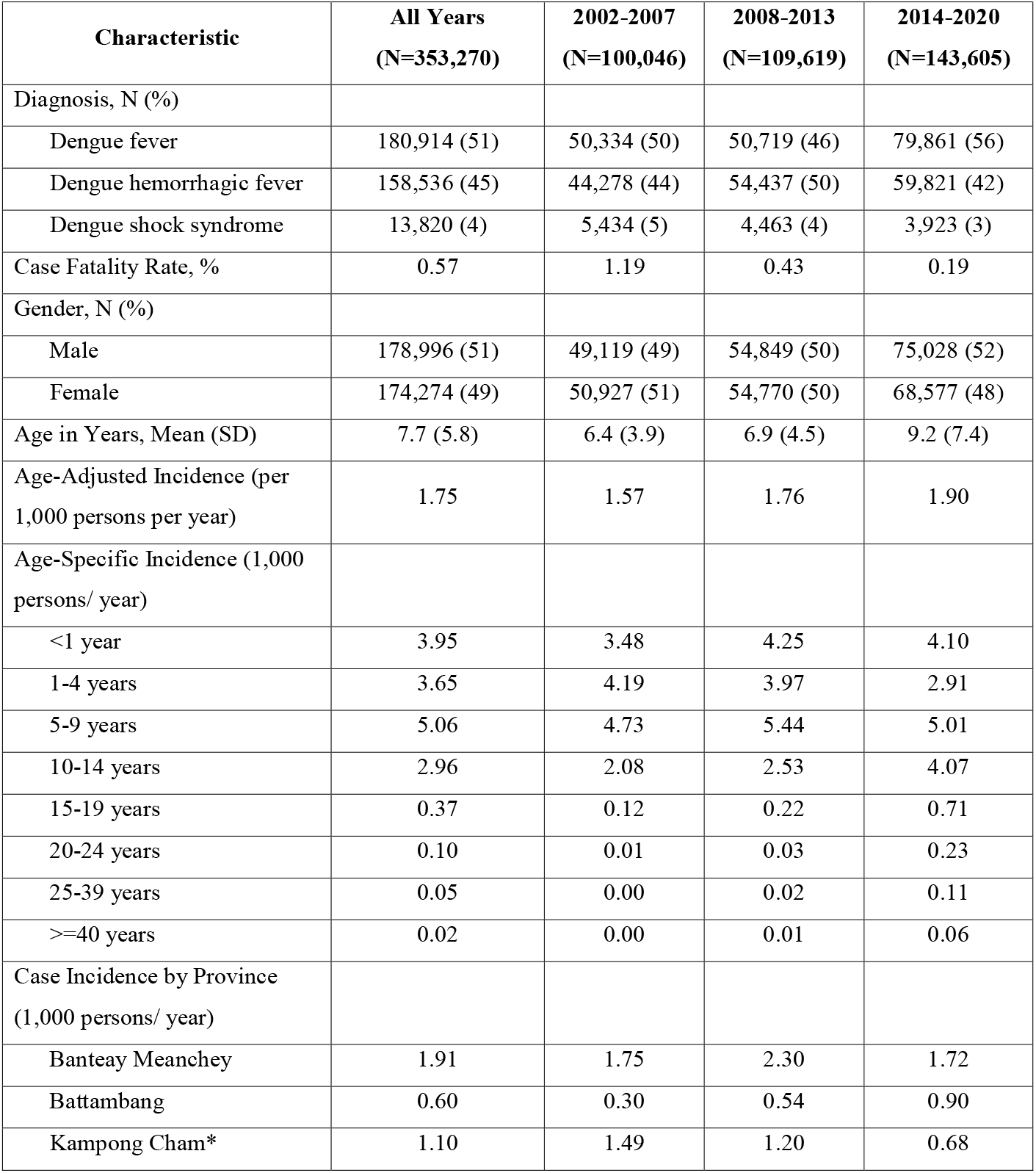
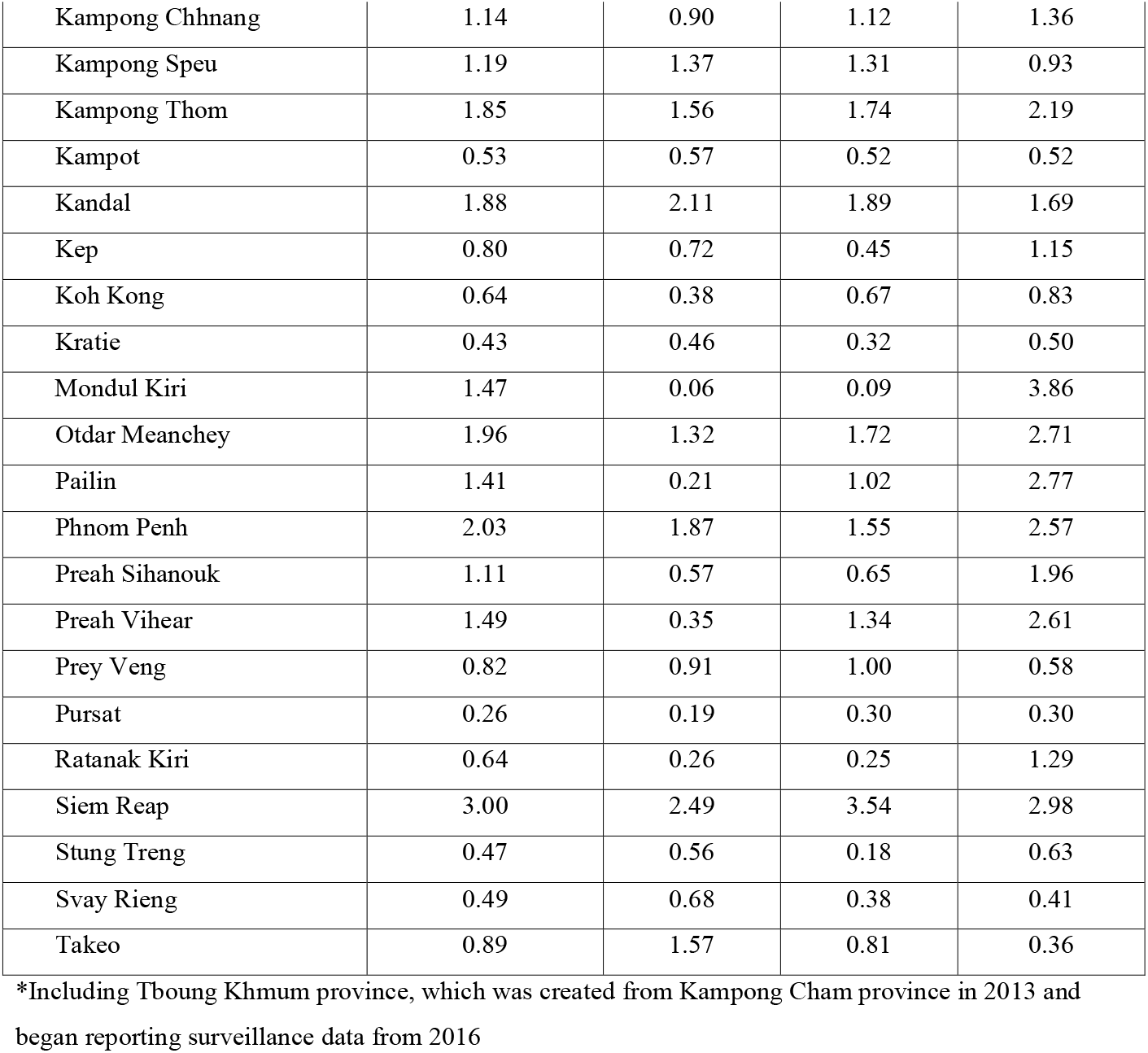
Demographic and Geographic Characteristics of Dengue Cases in Cambodia, 2002-2020.

| **Characteristic** | **All Years**<br>**(N=353,270)** | **2002-2007**<br>**(N=100,046)** | **2008-2013**<br>**(N=109,619)** | **2014-2020**<br>**(N=143,605)** |
| --- | --- | --- | --- | --- |
| Diagnosis, N (%) |  |  |  |  |
| Dengue fever | 180,914 (51) | 50,334 (50) | 50,719 (46) | 79,861 (56) |
| Dengue hemorrhagic fever | 158,536 (45) | 44,278 (44) | 54,437 (50) | 59,821 (42) |
| Dengue shock syndrome | 13,820 (4) | 5,434 (5) | 4,463 (4) | 3,923 (3) |
| Case Fatality Rate, % | 0.57 | 1.19 | 0.43 | 0.19 |
| Gender, N (%) |  |  |  |  |
| Male | 178,996 (51) | 49,119 (49) | 54,849 (50) | 75,028 (52) |
| Female | 174,274 (49) | 50,927 (51) | 54,770 (50) | 68,577 (48) |
| Age in Years, Mean (SD) | 7.7 (5.8) | 6.4 (3.9) | 6.9 (4.5) | 9.2 (7.4) |
| Age-Adjusted Incidence (per 1,000 persons per year) | 1.75 | 1.57 | 1.76 | 1.90 |
| Age-Specific Incidence (1,000 persons/ year) |  |  |  |  |
| <1 year | 3.95 | 3.48 | 4.25 | 4.10 |
| 1-4 years | 3.65 | 4.19 | 3.97 | 2.91 |
| 5-9 years | 5.06 | 4.73 | 5.44 | 5.01 |
| 10-14 years | 2.96 | 2.08 | 2.53 | 4.07 |
| 15-19 years | 0.37 | 0.12 | 0.22 | 0.71 |
| 20-24 years | 0.10 | 0.01 | 0.03 | 0.23 |
| 25-39 years | 0.05 | 0.00 | 0.02 | 0.11 |
| >=40 years | 0.02 | 0.00 | 0.01 | 0.06 |
| Case Incidence by Province (1,000 persons/ year) |  |  |  |  |
| Banteay Meanchey | 1.91 | 1.75 | 2.30 | 1.72 |
| Battambang | 0.60 | 0.30 | 0.54 | 0.90 |
| Kampong Cham* | 1.10 | 1.49 | 1.20 | 0.68 |
| Kampong Chhnang | 1.14 | 0.90 | 1.12 | 1.36 |
| Kampong Speu | 1.19 | 1.37 | 1.31 | 0.93 |
| Kampong Thom | 1.85 | 1.56 | 1.74 | 2.19 |
| Kampot | 0.53 | 0.57 | 0.52 | 0.52 |
| Kandal | 1.88 | 2.11 | 1.89 | 1.69 |
| Kep | 0.80 | 0.72 | 0.45 | 1.15 |
| Koh Kong | 0.64 | 0.38 | 0.67 | 0.83 |
| Kratie | 0.43 | 0.46 | 0.32 | 0.50 |
| Mondul Kiri | 1.47 | 0.06 | 0.09 | 3.86 |
| Otdar Meanchey | 1.96 | 1.32 | 1.72 | 2.71 |
| Pailin | 1.41 | 0.21 | 1.02 | 2.77 |
| Phnom Penh | 2.03 | 1.87 | 1.55 | 2.57 |
| Preah Sihanouk | 1.11 | 0.57 | 0.65 | 1.96 |
| Preah Vihear | 1.49 | 0.35 | 1.34 | 2.61 |
| Prey Veng | 0.82 | 0.91 | 1.00 | 0.58 |
| Pursat | 0.26 | 0.19 | 0.30 | 0.30 |
| Ratanak Kiri | 0.64 | 0.26 | 0.25 | 1.29 |
| Siem Reap | 3.00 | 2.49 | 3.54 | 2.98 |
| Stung Treng | 0.47 | 0.56 | 0.18 | 0.63 |
| Svay Rieng | 0.49 | 0.68 | 0.38 | 0.41 |
| Takeo | 0.89 | 1.57 | 0.81 | 0.36 |

\*Including Tboung Khmum province, which was created from Kampong Cham province in 2013 and began reporting surveillance data from 2016

**Figure 1.**
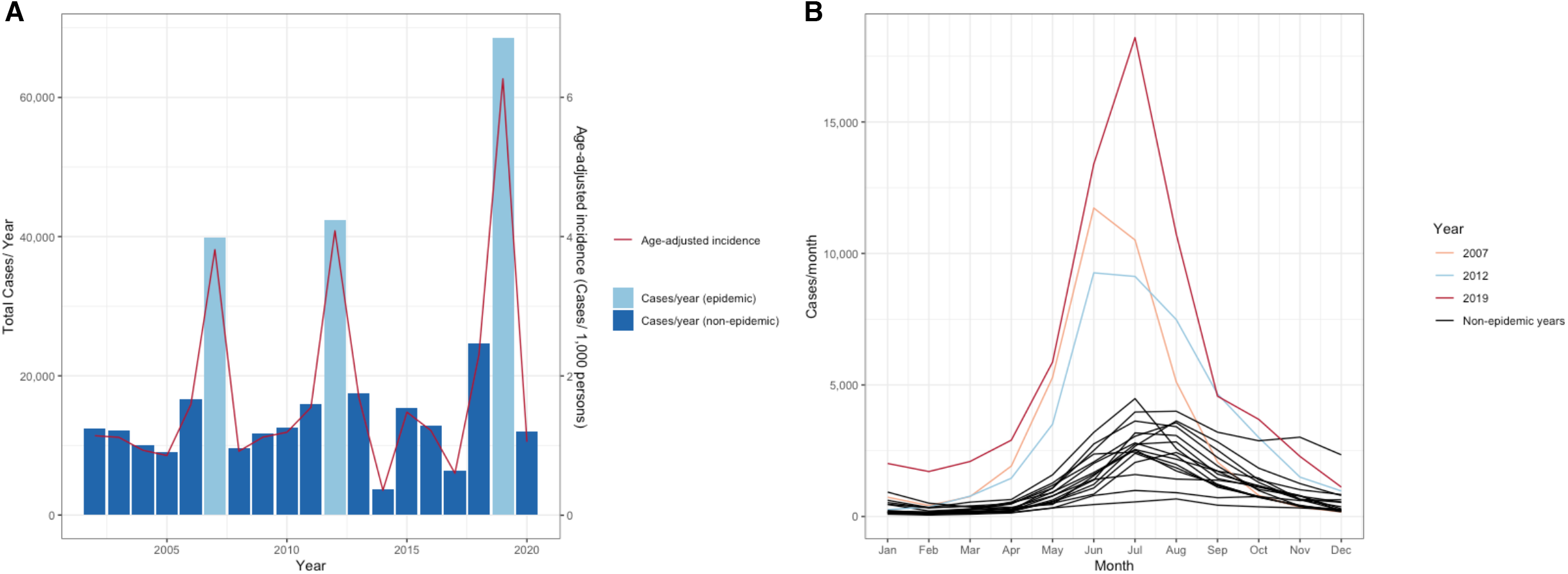
A) Annual dengue incidence in Cambodia, 2002-2020. Absolute dengue case numbers represented by blue bars on the left y-axis, case incidence (cases/ 1,000 persons) represented by the red line on the right y-axis. B) Monthly dengue case numbers in Cambodia, by year (2002-2020), with colors corresponding to specific epidemic years vs. non-epidemic years, as indicated by legend.

**Figure 2.**
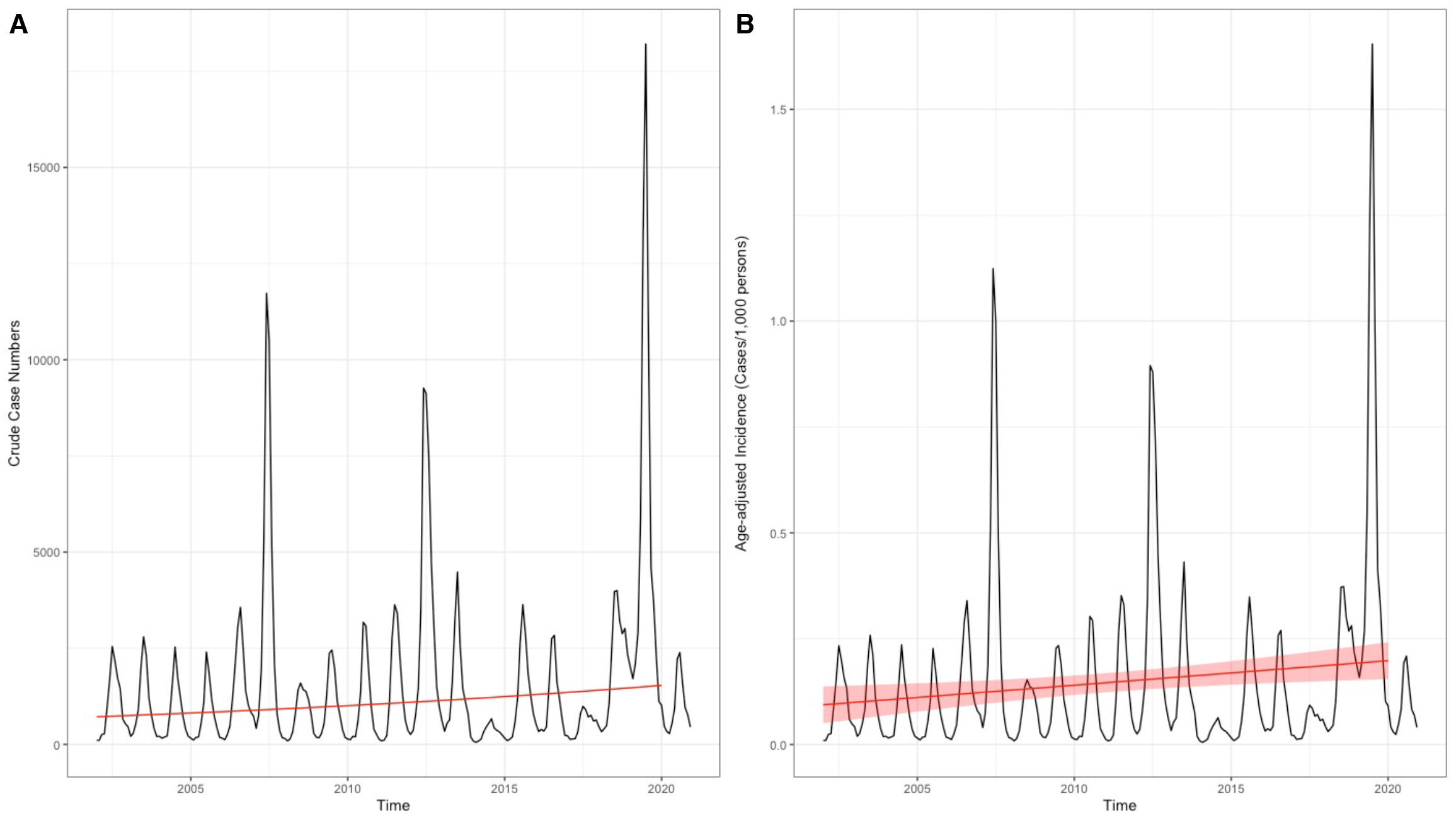
Generalized additive models (GAM) fitted to A) crude dengue case numbers and B) age-adjusted dengue case incidence in Cambodia from 2002 and 2020. Black lines depict crude cases (A) and age-adjusted incidence (B), respectively, while red line gives GAM projections excluding the effect of month. Translucent red shading corresponds to 95% confidence intervals by standard error.

### Changes in Dengue Case Characteristics Over Time

Dengue cases were evenly distributed by gender (51% male, 49% female), and mean age of infected individuals across the entire period was 7.7 years (SD 5.8). In a generalized additive model, predicted mean age of infected individuals increased significantly from 5.8 years (SE 0.3) in 2002 to 9.1 years (SE 0.4) in 2020 (slope = 0.18, SE = 0.0088, p <0.001; Figure 3A and Supplementary Table 1). The trend of increasing age with time was sustained within different case phenotypes (dengue fever: slope = 0.17, SE = 0.011, p <0.001; dengue hemorrhagic fever: slope = 0.20, SE = 0.0095, p <0.001; dengue shock syndrome: slope = 0.13, SE = 0.012, p <0.001; Supplementary Figure 3). DF represented the majority (51%) of overall cases across the entire study period, with DHF and DSS representing 45% and 4%, respectively. Average annual case fatality rate was 0.57% (SD 0.48). Generalized additive models demonstrated decreasing proportions of severe dengue among overall cases (DHF: 47.8% in 2002 to 42.6% in 2020, slope = -0.012, SE = 0.00062, p <0.001; DSS: 6.7% in 2002 to 2.3% in 2020, slope = -0.061, SE = 0.0016, p <0.001), and decreasing case fatality rates (1.77% in 2002 to 0.10% in 2020, slope = - 0.16, SE = 0.0050, p <0.001) from 2002 to 2020 (Figure 3B-D and Supplementary Table 1). Dengue serotype-specific RT-PCR was performed on a monthly basis in a subset of cases from sentinel sites. Across all years, DENV-2 was the most prevalent serotype (37.1%) followed by DENV-1 (32.2%), DENV-3 (22.6%), and DENV-4 (8.1%). There was significant yearly variation: the 2007 major epidemic was predominantly driven by DENV-3, while subsequent years saw alternating DENV-1 and DENV-2 predominance including the 2012 epidemic driven by DENV-1 and the 2019 epidemic driven by a mix of DENV-1 and DENV-2 (Supplementary Figure 3).

**Figure 3.**
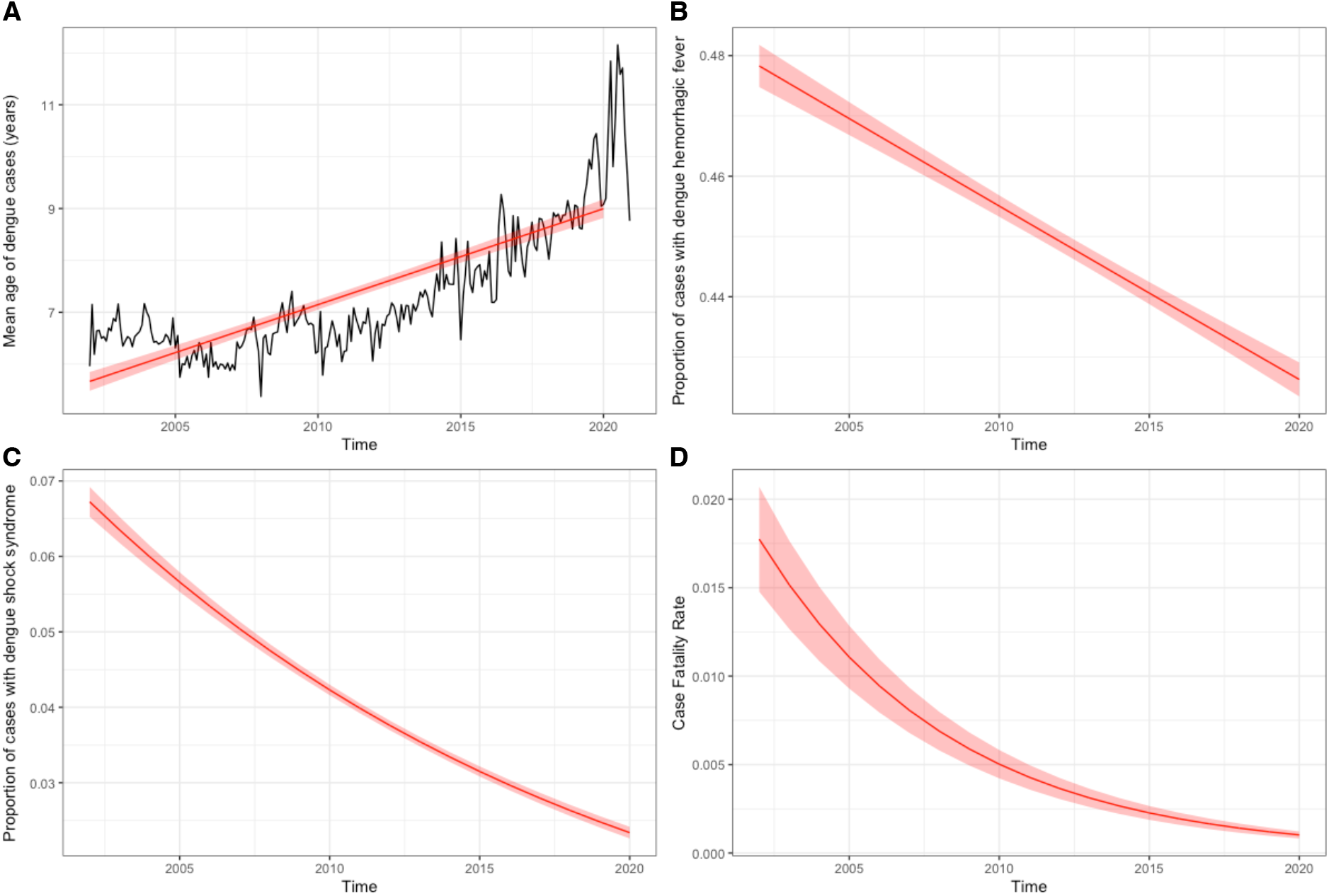
Generalized additive models of A) mean age of infected individuals, B) proportion of dengue hemorrhagic fever, C) proportion of dengue shock syndrome, and D) case fatality rate for all dengue cases in Cambodia from 2002 to 2020. Black line depicts mean age of infected individuals by month (A). Red lines depict GAM projections excluding the effect of month. Translucent shading corresponding to 95% confidence intervals by standard error.

### Comparisons of Data from National Surveillance Data and a Longitudinal Cohort Study

In July 2018, 771 children aged 2 to 9 years living in the province of Kampong Speu were enrolled in a community-based pediatric cohort study.^18^ Between July 2018-April 2020, a total of 51 clinically apparent, laboratory-confirmed dengue cases were identified (average monthly incidence (*P*_*A*_) of 3.0 per 1,000 persons (range: 0-22.0)) (Table 2). During the same period, national surveillance captured an average monthly incidence (*P*_*N*_) of 0.6 per 1,000 persons (range: 0.01-2.4) in the same age group and in the same province. The average case incidence ratio of *P*_*A*_*:P*_*N*_ was calculated to give an expansion factor (*EF*_*1*_) of 5.0 (95% CI 0.2-26.5). Among 597 children identified as immunologically naïve to dengue infection based on PRNT data at baseline assessment, there were a total of 148 cases of clinically inapparent seroconversions identified during July 2018-April 2020. The cumulative incidence of all dengue cases (*P*_*T*_; including both clinically apparent and inapparent cases) in the cohort was calculated at each 6-monthly study interval (average 6-monthly cumulative incidence 135.9 cases per 1,000 persons, range: 44.7-239.4) and compared to the corresponding 6-monthly cumulative incidence of apparent dengue cases detected by national surveillance (*P*_*N6*_; average 4.4 cases per 1,000 persons, range: 2.3-8.4); the ratio of *P*_*T*_*:P*_*N6*_ gave an expansion factor (*EF*_*2*_) of 33.6 (range: 18.7-53.7).

**Table 2:**
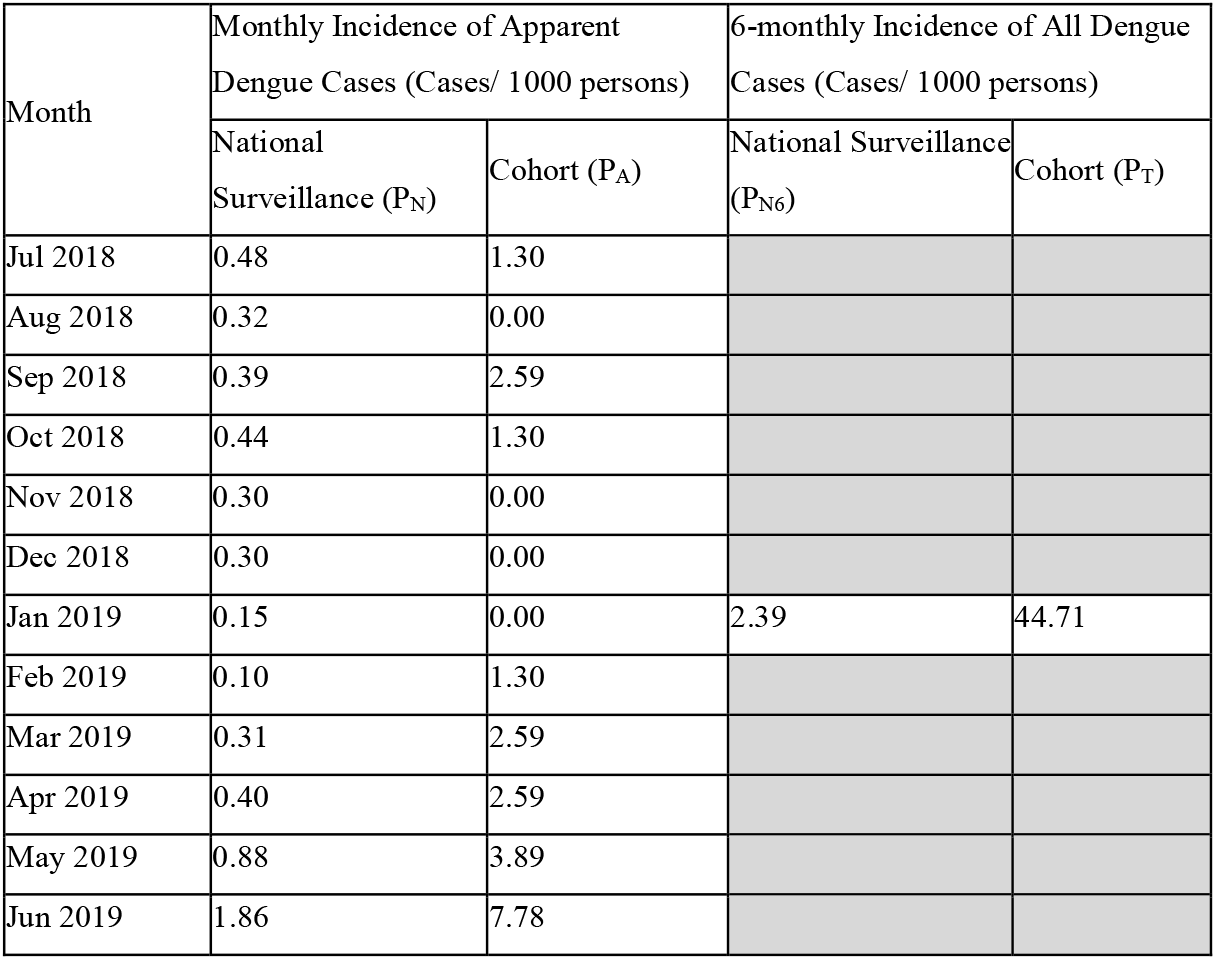
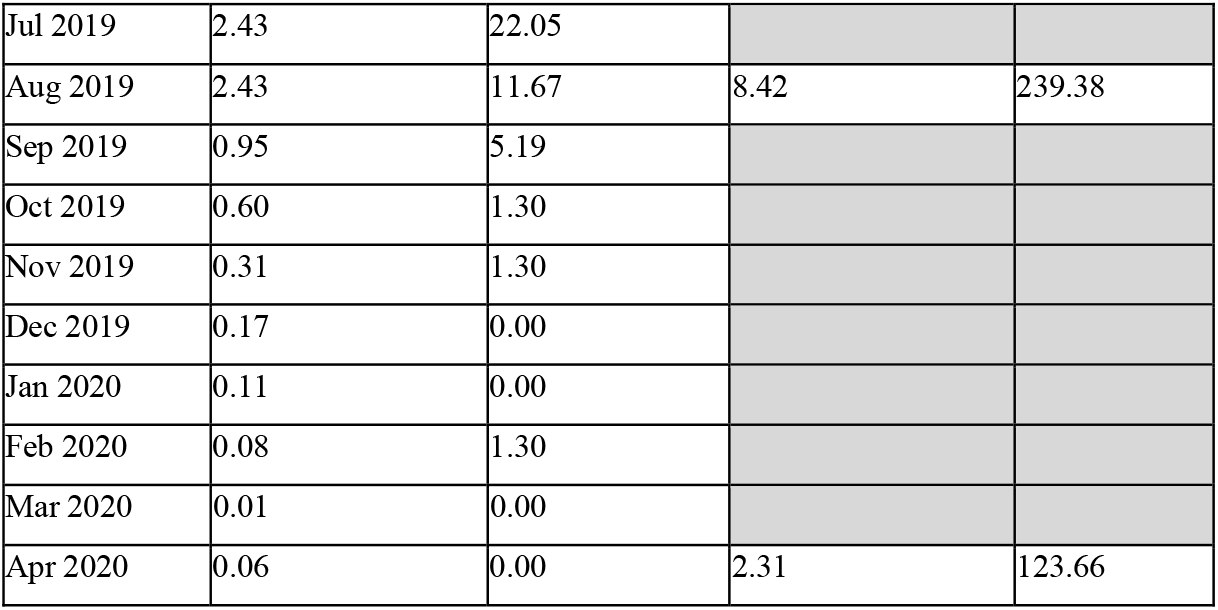
Comparison of dengue case incidence in Kampong Speu province from cohort and national data sets, July 2018-April 2020

## Discussion

Cambodia is in a dramatic state of change with accelerated rural-to-urban migration, climate change, land-use transformation, infrastructure development, and demographic and socio-economic shifts within its population. The dynamic milieu affects transmissibility of vector-borne disease and poses challenges to accurate modeling of dengue burden in this hyper-endemic country. Data from 19 years of national dengue surveillance in Cambodia show a greater than two-fold increase in dengue from 2002-2020, and a shift in the bulk of disease burden to older pediatric age groups. When compared with data from a local longitudinal pediatric cohort, national surveillance appears to under-estimate incidence by 5.0-fold for clinically apparent cases and 33.6-fold for both apparent and inapparent cases in Kampong Speu province.

Dengue is also increasing in other areas of Southeast Asia.^2,21^ Understanding the root causes of disease expansion can be challenging in countries where national surveillance is passive and inconsistent. While changes in surveillance practices could contribute to an apparent increase in cases, our comparison of national data with local cohort surveillance in 2018-2020 demonstrated comparable underreporting of dengue burden with that identified in previous work from earlier years.^13,14^ If we assume that national dengue reporting systems in Cambodia have remained fixed through time, possible explanations for the observed increase in dengue cases from 2002-2020 include: i) a steady increase in susceptible hosts, such as due to population growth (though we note that because dengue incidence increased as well, changes in population size alone cannot explain the observed rise), and ii) an increase in transmission intensity, such as due to increasing population density, a growing vector pool, altered vector/host behavior leading to heightened exposure, or evolving viral pathogenicity.^22^ Most likely, there are multifold contributors, and a multifaceted approach is needed to reduce transmission intensity.

We noted a concurrent increase in mean age of dengue infection in Cambodia between 2002 and 2020, predominantly driven by an increase in age-specific incidence in the 10-14 year age group. This effect may be underestimated due to poor clinical recognition of dengue cases in non-pediatric populations in Cambodia.^23^ The shift of dengue to older populations has been described in other SEA countries and attributed to demographic transitions resulting from industrialization.^9,24,25^ However, changes in the age distribution of the overall population alone are unlikely to account for this shift, as even within age strata we noted different behavior of age-specific incidence with time: for example, incidence was stagnant or decreasing for ages <10 years but increasing for ages 10-14 years. More likely, socioeconomic development leading to altered human behavior and vector habitats may have moved the host:vector interface away from the home and into public spaces such as schools and workplaces. Dengue has been classically described as a disease of the young, but recent trends may indicate the need for a paradigm shift. Recognizing the changing demographic of dengue, it is critical that interventions such as vaccination be planned using accurate age-specific data to identify appropriate target populations.

Disease under-detection is a problem in most dengue-endemic countries and various modeling methods have been proposed to address the issue.^6,7,26^ Two previous studies aimed to measure the degree of under-estimation of clinically apparent dengue in Kampong Cham province, Cambodia, between 2006 and 2008. They found 9.1-fold^13^ and 3.9 to 29.0-fold (variation by year)^14^ more symptomatic dengue cases detected with active surveillance of pediatric cohorts compared to national data, respectively. Neither study examined inapparent infection. Here, we applied data from a pediatric cohort in Kampong Speu province and found a 5.0-fold higher rate of clinically apparent dengue in cohort versus national data between 2018 and 2020. Varying rates of dengue and quality of healthcare infrastructure among provinces and over time are among multiple factors affecting surveillance fidelity – this is reflected by the wide variation of expansion factors reported here and elsewhere.^6^ An additional finding from our cohort was the high incidence of clinically inapparent dengue, accounting for an additional 6-fold underestimation of total case numbers. Ultimately, accurate capture of both clinically apparent and inapparent dengue is needed, for instance, to identify areas with high transmission that could benefit from vector control or populations with monotypic immunity for vaccination. Febrile and serologic surveillance should occur in tandem to inform public health interventions.^26^

Dengue control is a global health priority.^1^ Fortunately, despite increasing dengue incidence in Cambodia over the past two decades, the proportion of severe manifestations of dengue and case fatality rates have declined. This is likely due to socioeconomic progress leading to improving overall population health (increased access to care, fewer co-morbidities). Additionally, the National Dengue Control Program has focused on intensive education and in-service campaigns for nurses and doctors over the last few years,^27^ and has urged treatment of dengue patients within the decentralized national health system instead of at unregulated private practices.^16^ These improvements may have enabled earlier recognition and appropriate management of cases, and are overall an encouraging sign that public health efforts in Cambodia are producing positive results.

This study has several limitations. Effects of climate, migration, changing surveillance practices, advances in clinical knowledge, improvements in health infrastructure, and periodic boosting of public and provider awareness could not be accounted for given inconsistent collection of these data over the 19-year period. The last comprehensive assessment of dengue epidemiology in Cambodia included an evaluation of vector control, but this analysis could not be repeated due to limited updated data.^12^

## Conclusion

The incidence of dengue in Cambodia has increased significantly over the past two decades and disease burden is shifting to older pediatric populations. True burden continues to be under-estimated, although there have been notable successes of national programs reflected by a reduction in cases of severe dengue and case fatality rates. Future interventions such as vaccine campaigns or vector control will need to account for under-estimation and shifting demographics to target susceptible populations at the appropriate scale.

## Data Availability

All data produced in the present study are available upon reasonable request to the authors.

## Acknowledgements

The authors would like to acknowledge the staff at the National Dengue Control Program and the National Center of Parasitology, Entomology, and Malaria Control in Cambodia for their contributions to dengue surveillance, control, and reporting.

## Data Availability Statement

All data produced in the present study are available upon reasonable request to the authors.

## References

1. World Health Organization. Dengue and severe dengue. World Health Organization, Geneva, 2021. Accessed August 31, 2021. https://www.who.int/news-room/fact-sheets/detail/dengue-and-severe-dengue

2. Simmons CP, Farrar JJ, Nguyen van VC, Wills B. Dengue. N Engl J Med. 2012;366(15):1423–1432. doi:10.1056/NEJMra1110265

3. Sridhar S, Luedtke A, Langevin E, et al. Effect of Dengue Serostatus on Dengue Vaccine Safety and Efficacy. New England Journal of Medicine. 2018;379(4):327–340. doi:10.1056/NEJMoa1800820

4. Biswal S, Borja-Tabora C, Martinez Vargas L, et al. Efficacy of a tetravalent dengue vaccine in healthy children aged 4-16 years: a randomised, placebo-controlled, phase 3 trial. Lancet. 2020;395(10234):1423–1433. doi:10.1016/S0140-6736(20)30414-1

5. Kallas EG, Precioso AR, Palacios R, et al. Safety and immunogenicity of the tetravalent, live-attenuated dengue vaccine Butantan-DV in adults in Brazil: a two-step, double-blind, randomised placebo-controlled phase 2 trial. Lancet Infect Dis. 2020;20(7):839–850. doi:10.1016/S1473-3099(20)30023-2

6. Undurraga EA, Halasa YA, Shepard DS. Use of expansion factors to estimate the burden of dengue in Southeast Asia: a systematic analysis. PLoS Negl Trop Dis. 2013;7(2):e2056. doi:10.1371/journal.pntd.0002056

7. Stanaway JD, Shepard DS, Undurraga EA, et al. The global burden of dengue: an analysis from the Global Burden of Disease Study 2013. Lancet Infect Dis. 2016;16(6):712–723. doi:10.1016/S1473-3099(16)00026-8

8. Grange L, Simon-Loriere E, Sakuntabhai A, Gresh L, Paul R, Harris E. Epidemiological Risk Factors Associated with High Global Frequency of Inapparent Dengue Virus Infections. Front Immunol. 2014;5:280. doi:10.3389/fimmu.2014.00280

9. Katzelnick LC, Ben-Shachar R, Mercado JC, et al. Dynamics and determinants of the force of infection of dengue virus from 1994 to 2015 in Managua, Nicaragua. Proc Natl Acad Sci U S A. 2018;115(42):10762–10767. doi:10.1073/pnas.1809253115

10. Lopez AL, Adams C, Ylade M, et al. Determining dengue virus serostatus by indirect IgG ELISA compared with focus reduction neutralisation test in children in Cebu, Philippines: a prospective population-based study. Lancet Glob Health. 2021;9(1):e44–e51. doi:10.1016/S2214-109X(20)30392-2

11. Reiner RC, Stoddard ST, Forshey BM, et al. Time-varying, serotype-specific force of infection of dengue virus. Proc Natl Acad Sci U S A. 2014;111(26):E2694–2702. doi:10.1073/pnas.1314933111

12. Huy R, Buchy P, Conan A, et al. National dengue surveillance in Cambodia 1980–2008: epidemiological and virological trends and the impact of vector control. Bulletin of the World Health Organization. 2010;88(9):650. doi:10.2471/BLT.09.073908

13. Wichmann O, Yoon IK, Vong S, et al. Dengue in Thailand and Cambodia: An Assessment of the Degree of Underrecognized Disease Burden Based on Reported Cases. PLoS Negl Trop Dis. 2011;5(3):e996. doi:10.1371/journal.pntd.0000996

14. Vong S, Goyet S, Ly S, et al. Under-recognition and reporting of dengue in Cambodia: a capture-recapture analysis of the National Dengue Surveillance System. Epidemiol Infect. 2012;140(3):491–499. doi:10.1017/S0950268811001191

15. Ou TP, Yun C, Auerswald H, et al. Improved detection of dengue and Zika viruses using multiplex RT-qPCR assays. Journal of Virological Methods. 2020;282:113862. doi:10.1016/j.jviromet.2020.113862

16. Cambodian Ministry of Health. National Guideline for Clinical Management of Dengue (Version 3). Published online 2018. Accessed October 28, 2021. https://niph.org.kh/niph/uploads/library/pdf/GL055_National_guideline_for_ClM_Dengue.pdf

17. The World Bank. World Development Indicators. Accessed October 29, 2021. https://databank.worldbank.org/metadataglossary/2/series/SP.POP.GROW

18. Manning JE, Chea S, Parker DM, et al. Development of inapparent dengue associated with increased antibody levels to Aedes aegypti salivary proteins: a longitudinal dengue cohort in Cambodia. J Infect Dis. Published online October 27, 2021:jiab541. doi:10.1093/infdis/jiab541

19. World Health Organization Regional Office for the Western Pacific. Dengue Situation Updates. Published online 2020. http://iris.wpro.who.int/handle/10665.1/14461

20. Cousien A, Ledien J, Souv K, et al. Predicting Dengue Outbreaks in Cambodia. Emerg Infect Dis. 2019;25(12):2281–2283. doi:10.3201/eid2512.181193

21. Du M, Jing W, Liu M, Liu J. The Global Trends and Regional Differences in Incidence of Dengue Infection from 1990 to 2019: An Analysis from the Global Burden of Disease Study 2019. Infect Dis Ther. 2021;10(3):1625–1643. doi:10.1007/s40121-021-00470-2

22. Katzelnick LC, Coello Escoto A, Huang AT, et al. Antigenic evolution of dengue viruses over 20 years. Science. 2021;374(6570):999–1004. doi:10.1126/science.abk0058

23. Bohl Jennifer A., Lay Sreyngim, Chea Sophana, et al. Discovering disease-causing pathogens in resource-scarce Southeast Asia using a global metagenomic pathogen monitoring system. Proceedings of the National Academy of Sciences. 2022;119(11):e2115285119. doi:10.1073/pnas.2115285119

24. Cummings DAT, Iamsirithaworn S, Lessler JT, et al. The Impact of the Demographic Transition on Dengue in Thailand: Insights from a Statistical Analysis and Mathematical Modeling. PLoS Med. 2009;6(9):e1000139. doi:10.1371/journal.pmed.1000139

25. Rodríguez-Barraquer I, Buathong R, Iamsirithaworn S, et al. Revisiting Rayong: Shifting Seroprofiles of Dengue in Thailand and Their Implications for Transmission and Control. American Journal of Epidemiology. 2014;179(3):353–360. doi:10.1093/aje/kwt256

26. Bhatt S, Gething PW, Brady OJ, et al. The global distribution and burden of dengue. Nature. 2013;496(7446):504–507. doi:10.1038/nature12060

27. Handel AS, Ayala EB, Borbor-Cordova MJ, et al. Knowledge, attitudes, and practices regarding dengue infection among public sector healthcare providers in Machala, Ecuador. Trop Dis Travel Med Vaccines. 2016;2:8. doi:10.1186/s40794-016-0024-y

28. Wood SN. Mgcv: GAMs and generalized ridge regression for R. R News. 2001;1(2):20–25.

