## Supplemental data for "Dengue in Cambodia 2002-2020: Cases, Characteristics and Capture by National Surveillance"

**Supplemental Text S1:**

Generalized additive models were fitted to time series of monthly dengue crude case numbers, age-adjusted case incidence, mean age of infected individuals, and case phenotype/ outcome from 2002-2020. The models included a fixed predictor of year and a monthly smoothing term (with the number of smoothing knots, k, fixed at 7, and incorporating a cyclic cubic smoothing spline).^28^ Final models took on the following construction:

gam(crude.case.numbers ~ year + s(month, k=7, bs="cc"), family="poisson")

gam(age.adjusted.incidence ~ year + s(month, k=7, bs="cc"), family="gaussian")

gam(mean.age ~ year + s(month, k=7, bs="cc"), family="gaussian")

gam(dengue.fever ~ year + s(month, k=7, bs="cc"), family="binomial")

gam(dengue.hemorrhagic.fever~ year + s(month, k=7, bs="cc"), family="binomial")

gam(dengue.shock.syndrome ~ year + s(month, k=7, bs="cc"), family="binomial")

gam(case.fatality ~ year + s(month, k=7, bs="cc"), family="binomial")

**Supplementary Table 1:** Output from fitted generalized additive models

|  | **Fixed effect (Year)** | | | | **Smooth term (Month)** | | | **R^2^** |
| --- | --- | --- | --- | --- | --- | --- | --- | --- |
| **Variable** | **Slope** | **Std. Error** | **z-value** | **P-value** | **edf** | **Chi square** | **P-value** |  |
| **Poisson/ Binomial Models** | | | | | | | | |
| Crude case numbers | 0.042 | 0.00031 | 135 | <2x10^-16^ | 4.96 | 183217 | <2x10^-16^ | 0.334 |
| Dengue fever | 0.021 | 0.00062 | 33.9 | <2x10^-16^ | 4.95 | 2330 | <2x10^-16^ | 0.0093 |
| Dengue hemorrhagic fever | -0.012 | 0.00062 | -18.9 | <2x10^-16^ | 4.95 | 1977 | <2x10^-16^ | 0.0063 |
| Dengue shock syndrome | -0.061 | 0.0016 | -38.6 | <2x10^-16^ | 4.24 | 161.9 | <2x10^-16^ | 0.0051 |
| Case fatality | -0.16 | 0.0050 | -32.1 | <2x10^-16^ | 3.84 | 63.5 | <2x10^-16^ | 0.0036 |
| **Gaussian Models** | | | | | | | | |
| **Variable** | **Slope** | **Std. Error** | **t-value** | **P-value** | **edf** | **F-statistic** | **P-value** | **R^2^** |
| Age-adjusted incidence | 0.0058 | 0.0021 | 2.8 | 0.0064 | 4.04 | 20.7 | <2x10^-16^ | 0.325 |
| Mean age (all cases types) | 0.18 | 0.0088 | 21.1 | <2x10^-16^ | 1.60 | 1.16 | 0.018 | 0.665 |
| Mean age (dengue fever) | 0.17 | 0.011 | 15.3 | <2x10^-16^ | 1.88 | 2.47 | 0.0005 | 0.520 |
| Mean age (dengue hemorrhagic fever) | 0.20 | 0.0095 | 20.7 | <2x10^-16^ | 1.84 | 1.70 | 0.005 | 0.657 |
| Mean age (dengue shock syndrome) | 0.13 | 0.012 | 10.4 | <2x10^-16^ | 1x10^-8^ | 0 | 0.39 | 0.324 |

**Supplementary Figure 1:**

Dengue cases by year and province. Phnom Penh and Siem Reap provinces, where overall case incidence is highest among all provinces, and Kampong Cham and Kampong Speu provinces, from which surveillance cohorts have been applied to estimate expansion factors, are denoted by colored lines.

**Supplementary Figure 2:**

GAM models fitted to mean age of infected individuals with A) dengue fever, B), dengue hemorrhagic fever, and C) dengue shock syndrome in Cambodia from 2002 to 2020. Black lines depict mean age in years, red line gives annual GAM projections, excluding the effect of month. Translucent red shading corresponds to 95% confidence intervals by standard error.

**Supplementary Figure 3:** Dengue serotype prevalence by year, based on RT-PCR performed on samples from sentinel sites. Shaded bars correspond to the number of samples tested (left y-axis) while colored lines indicate proportion of samples by serotype (right y-axis), as indicated in the legend.
