## Supplementary figures and images for "Dengue in Cambodia 2002-2020: Cases, Characteristics and Capture by National Surveillance"

### Supplemental Figure 1

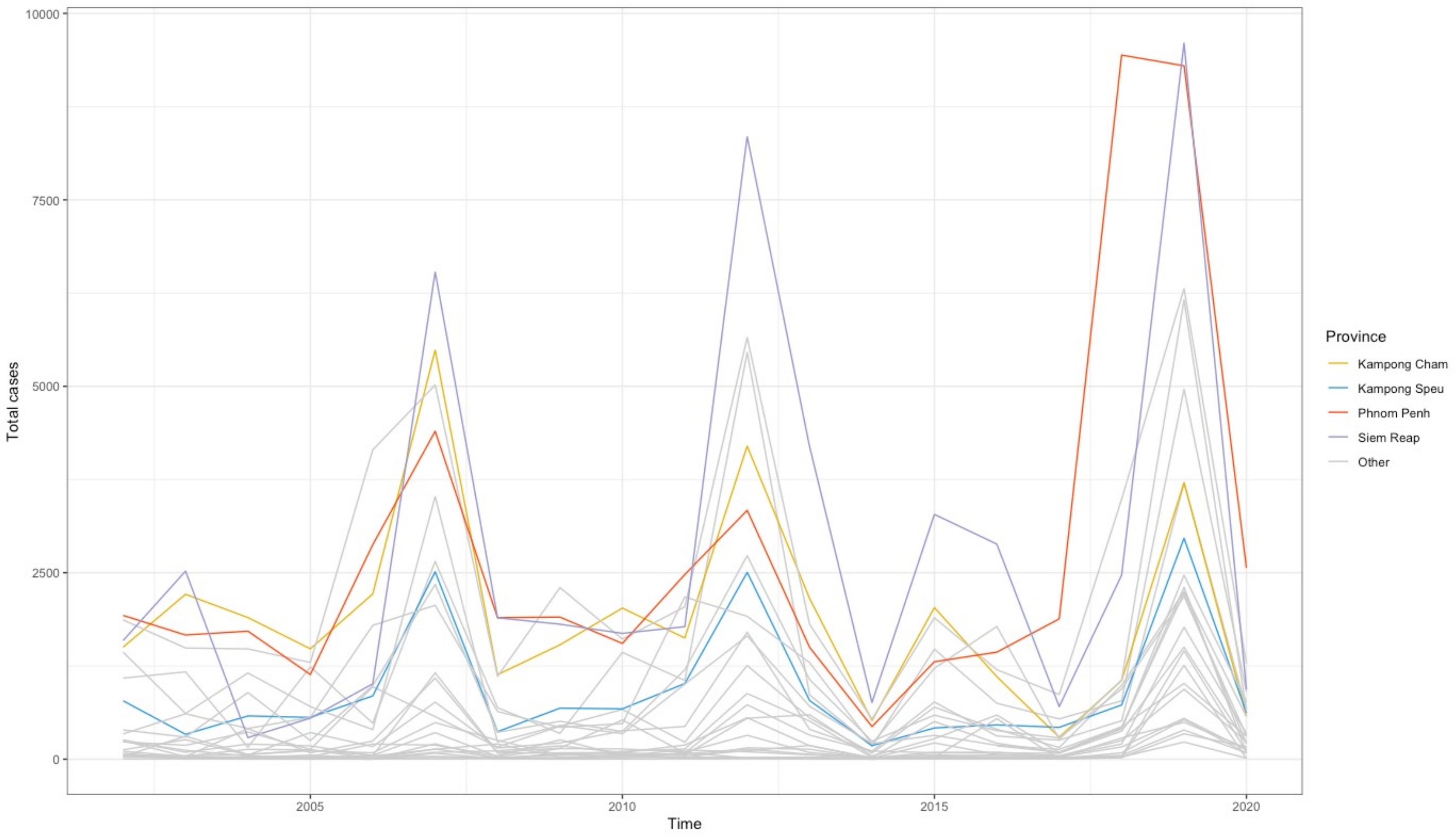

### Supplemental Figure 2

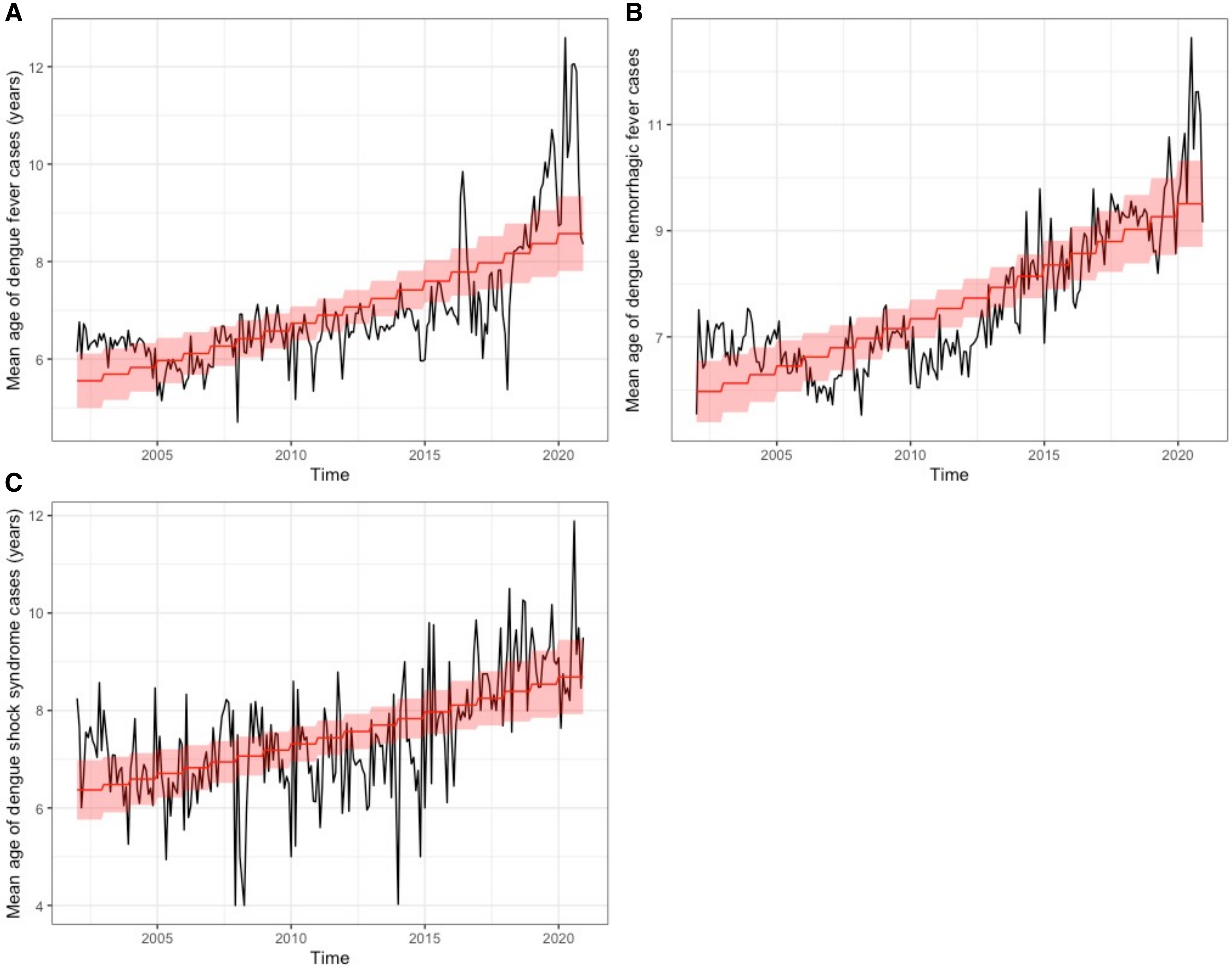

### Supplemental Figure 3

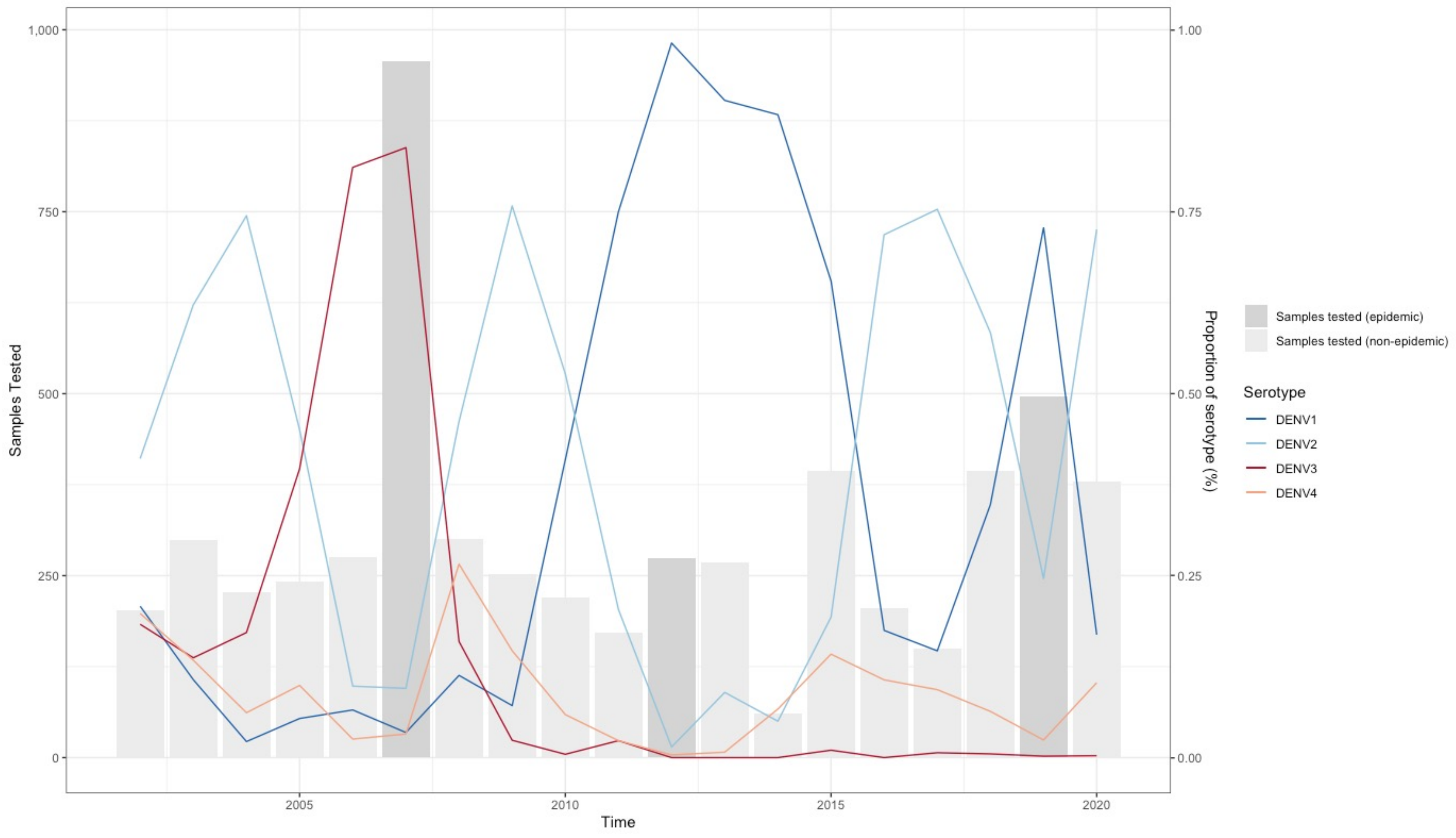
